# Hospital length of stay in a mixed Omicron and Delta epidemic in New South Wales, Australia

**DOI:** 10.1101/2022.03.16.22271361

**Authors:** Ruarai J Tobin, James G Wood, Duleepa Jayasundara, Grant Sara, James Walker, Genevieve E Martin, James M McCaw, Freya M Shearer, David J Price

## Abstract

**Aim:** To estimate the length of stay distributions of hospitalised COVID-19 cases during a mixed Omicron-Delta epidemic in New South Wales, Australia (16 Dec 2021 – 7 Feb 2022), and compare these to estimates produced over a Delta-only epidemic in the same population (1 Jul 2021 – 15 Dec 2022).

**Background:** The distribution of the duration that clinical cases of COVID-19 occupy hospital beds (the ‘length of stay’) is a key factor in determining how incident caseloads translate into health system burden as measured through ward and ICU occupancy.

**Results:** Using data on the hospital stays of 19,574 individuals, we performed a competing-risk survival analysis of COVID-19 clinical progression. During the mixed Omicron-Delta epidemic, we found that the mean length of stay for individuals who were discharged directly from ward without an ICU stay was, for age groups 0-39, 40-69 and 70+ respectively, 2.16 (95% CI: 2.12–2.21), 3.93 (95% CI: 3.78–4.07) and 7.61 days (95% CI: 7.31–8.01), compared to 3.60 (95% CI: 3.48–3.81), 5.78 (95% CI: 5.59–5.99) and 12.31 days (95% CI: 11.75–12.95) across the preceding Delta epidemic (15 Jul 2021 – 15 Dec 2021). We also considered data on the stays of individuals within the Hunter New England Local Health District, where it was reported that Omicron was the only circulating variant, and found mean ward-to-discharge length of stays of 2.05 (95% CI: 1.80–2.30), 2.92 (95% CI: 2.50–3.67) and 6.02 days (95% CI: 4.91–7.01) for the same age groups.

**Conclusions:** Hospital length of stay was substantially reduced across all clinical pathways during a mixed Omicron-Delta epidemic compared to a prior Delta epidemic. These changes in length of stay have contributed to lessened health system burden despite greatly increased infection burden and should be considered in future planning of response to the COVID-19 pandemic in Australia and internationally.

## Introduction

The state of New South Wales (NSW), Australia, employed a broadly successful suppression strategy against COVID-19 for the early period of the pandemic, with relatively low case incidence prior to an outbreak of the Delta variant in June 2021. This Delta epidemic grew substantially despite stepped escalation of public health and social measures, supported by intensive test-trace-isolate-quarantine measures. Cases totalled to 53,851 notifications across a two month period of August and September 2021, with a peak in hospital occupancy of 1,268 beds on 21 September. Subsequently, daily cases continued to decline, reaching a plateau in the low hundreds by late-October 2021 as the effective reproduction number stabilised below 1 — in part due to substantial vaccination uptake [1].

The Omicron variant was first identified in the New South Wales population on 28 November 2021 [2, 3, 4], with significant transmission including super-spreading events occurring in the following four weeks [5]. Notified COVID-19 cases continued to increase sharply, from 288 on 1 December 2021 up to 2,250 on 15 December. By the period of 10–24 January, Omicron represented 94% of sequenced cases in Australia [6, 3]. A total of 826,942 case notifications were made across a two month period from December 2021 through January 2022, which translated into a peak hospital occupancy of 2,943 reported on 25 January. Figure 1 shows case notifications and the reported ward and ICU occupancy through the study periods by age group. The peak hospital occupancy, whilst larger in absolute terms than the preceding Delta epidemic, indicates a relative reduction in the clinical demand resulting from the high number of cases during the Omicron epidemic.

**Figure 1:**
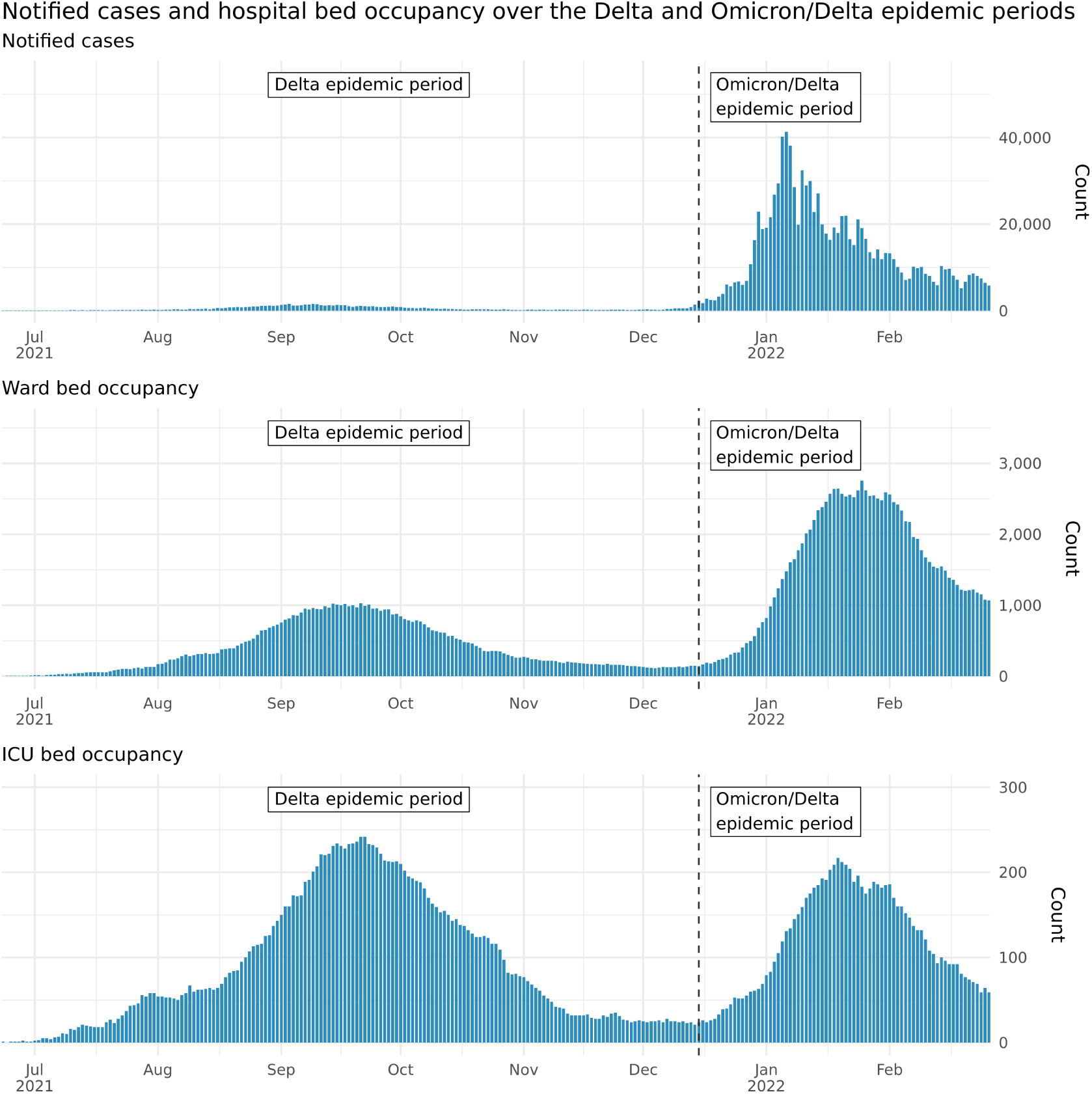
Number of COVID-19 cases by notification date and count of ward and ICU beds occupied by COVID-19 patients across the New South Wales hospital system. Case notification data is via Data.NSW and the NSW Ministry of Health [7, 8], hospital occupancy data via publicly reported figures collected by covid19data.com.au [9]. The number of notified cases may differ from counts reported at that date where later de-duplication of cases has been performed. Plotted data ranges from 1 July 2021 to 7 February 2022, with the dashed vertical line indicating the end date of the Delta epidemic period/start date of the mixed Omicron-Delta epidemic period for the purposes of this analysis.

We examined the length of stay distributions of hospitalised COVID-19 cases in NSW in two periods: the Delta only epidemic (1 July 2021 to 14 December 2021) and the mixed Omicron and Delta epidemic (15 December 2021 to 7 February 2022) to estimate the reduction in hospital length of stay during these periods. For the latter period, we further examined length of stay for a subset of cases hospitalised in the Hunter New England Local Health District of NSW where Omicron was reportedly the dominant, if not only, circulating variant. These time windows were chosen to characterise the outbreaks of Delta and Omicron variants in the absence of sufficient patient-level variant data. We performed an age-specific competing-risk survival analysis using a multi-state model of COVID-19 clinical progression, and report parameters for estimated length of stay distributions and corresponding summary statistics such that these results may be used to inform ongoing modelling and policy decisions.

## Methods

### Data

Episode level hospital stay data of COVID-19 patients was provided by the NSW Ministry of Health extracted from the routinely collected Admitted Patient Data Collection (APDC). Each datum consists of a singular episode within a ward with associated dates and times of admission and discharge. The date of symptom onset and age of each individual was known, but no information on vaccination status or infecting strain was available. In order to provide reliable estimates of the length of stay of patients, we performed several filtering steps to the data prior to fitting the multi-state model. Where an individual had been admitted to the intensive care unit (ICU), the initial and final known date and time were available in the data, alongside a total duration within ICU (which excluded any periods of ward stay between these initial and final known dates). Where the recorded duration in ICU was less than that indicated by the admission and discharge dates and times (which differed in only approximately 6% of admissions), we assumed that the additional time outside the ICU contributed to the post-ICU ward stay. This ensured – within the limitations of the compartmental model – that any ward stay that occurred during the period from a patient’s initial to final ICU date would not contribute to our estimates of ICU length of stay.

As an individual could have numerous episodes across multiple wards (e.g. representing transfer between different wards), episodes were concatenated to produce a single stay, where an individual would be assumed to be in hospital from the time of their earliest episode’s admission until the time of their latest episode’s discharge. At this stage, individuals were excluded from analysis where any two of their consecutive episodes were separated by at least 48 hours outside of the hospital system (n = 619, 1,442 and 85 for Delta, mixed-Omicron-Delta and HNE Omicron epidemic periods respectively; Figure 2). This filtering ensured that all cases included were valid under the single-stay assumption of the compartmental model.

**Figure 2:**
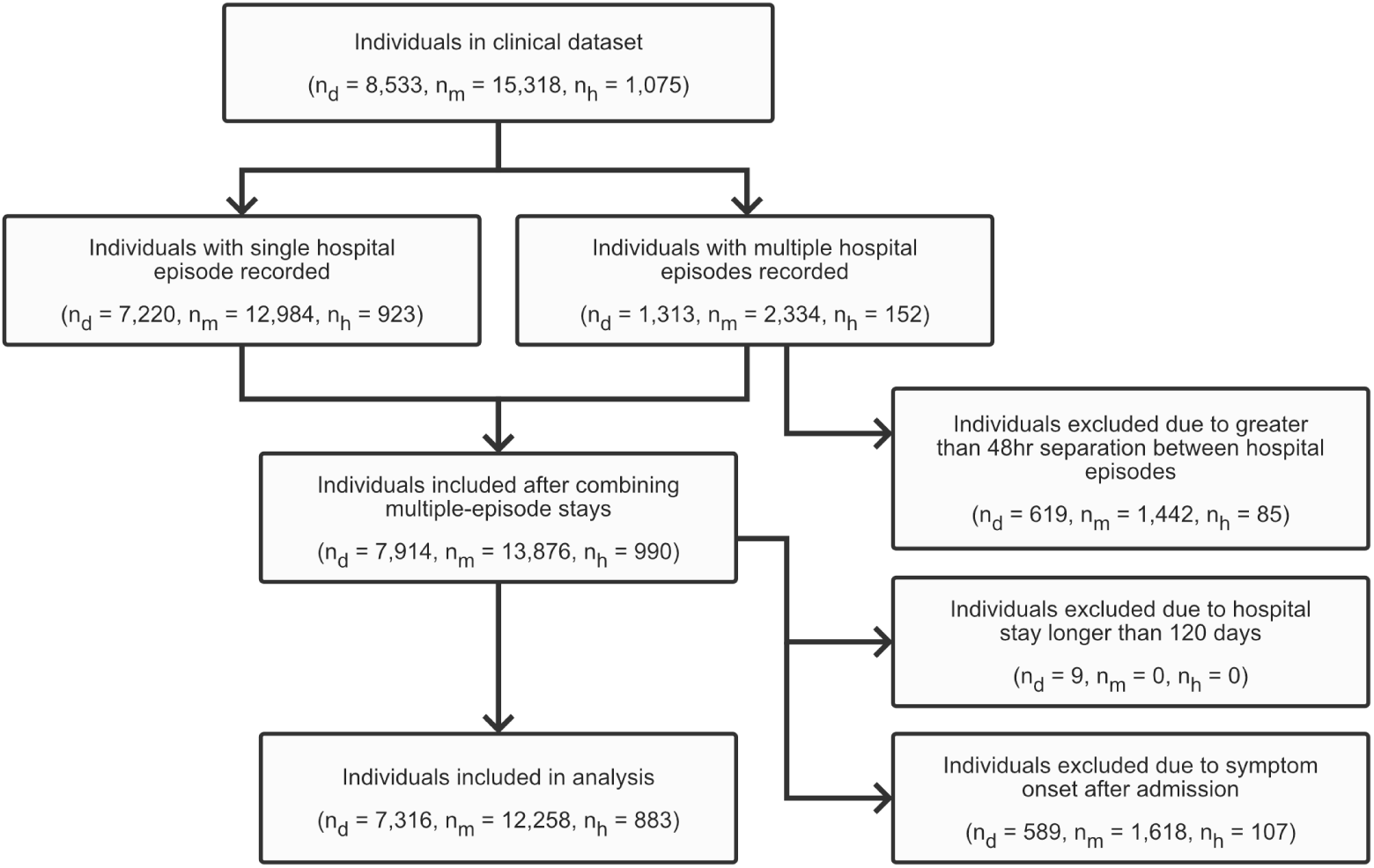
Data inclusion flowchart, where n_*d*_, n_*m*_ and n_*h*_ is the number of individuals in the Delta epidemic period, mixed-Omicron-Delta epidemic period and Omicron Hunter New England epidemic period analysis respectively. Note that the individuals included in the Omicron Hunter New England analysis are also included in the mixed-Omicron-Delta analysis.

Cases were removed from analysis where symptom onset was recorded to have occurred later than their earliest admission to hospital, given that that such infections were likely to have occurred within the hospital (n = 589, 1,618 and 107 for Delta, mixed-Omicron-Delta and HNE Omicron epidemic periods respectively; Figure 2). Although such within hospital infections are equally (if not more) likely to progress to a clinically severe state, the lack of differentiation in COVID-19 severity available to our analysis required the removal of these cases. A sensitivity analysis was performed to confirm that estimated lengths of stay were robust to this filtering assumption (Figure 9).

Individuals with a total hospital stay duration of greater than 120 days were removed from analysis (n = 9, 0 and 0 for Delta, mixed-Omicron-Delta and HNE Omicron epidemic periods respectively; Figure 2) as these were expected to be more likely to be incidental infections, with this assumption validated by examination of the ward and subward they were recorded as occupying. Furthermore, it was expected that such substantial stay durations (even in the case of them being true sequelae of infection) would have a disproportionate effect in the fitting of the parametric distributions to length of stay.

### Multi-state survival model

We performed an age-specific competing-risk survival analysis using a multi-state model of COVID-19 clinical progression (Figure 3). This multi-state model framework has been used for similarly characterising hospital demand for other patients infected with other SARS-CoV-2 variants [10, 11] and has been shown to produce the most reliable estimates of length of stay during an epidemic compared to other common survival approaches [12].

**Figure 3:**
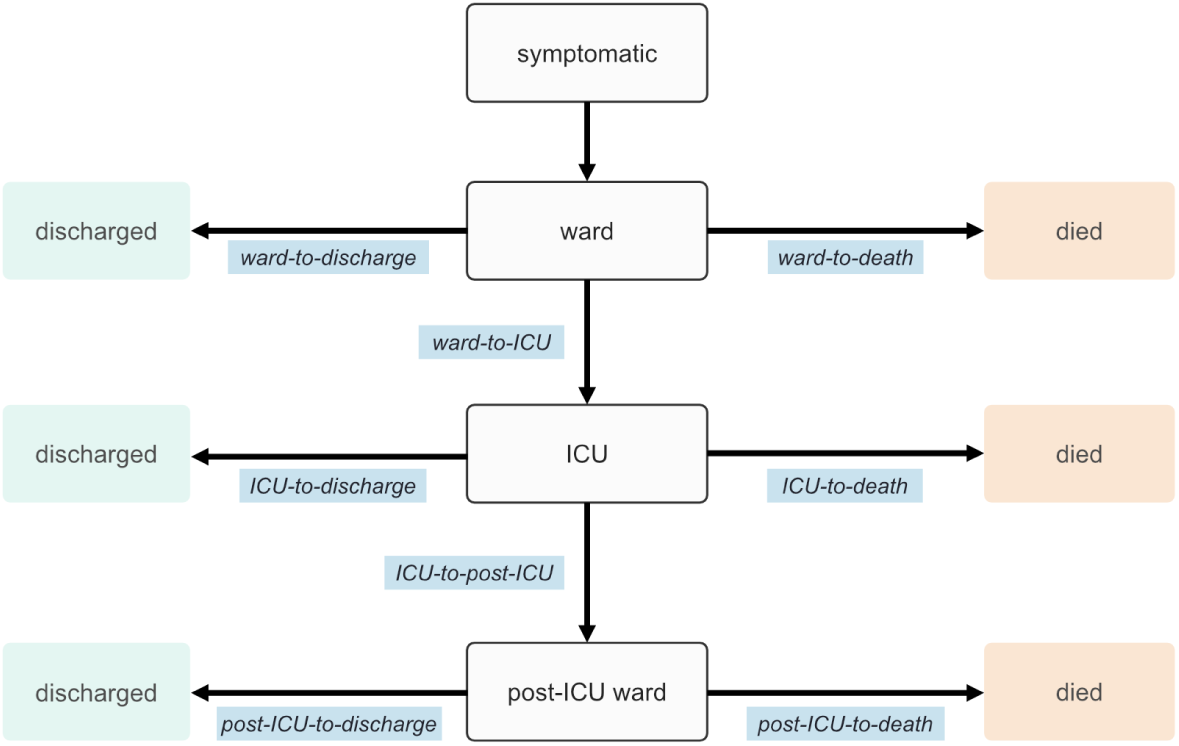
Compartmental model of COVID-19 clinical progression, with length of stay time distributions to be estimated across model transitions displayed in blue.

At the time of analysis, 11% of individuals in the mixed Omicron and Delta epidemic period data had not yet been observed to have an outcome (e.g. at the ward stage, individuals who have been admitted and not yet been discharged, transferred to ICU or died). If this censoring was not appropriately considered in the analysis, estimates would be biased down by an over-representation of individual’s with a shorter length of stay [12]. We considered each branching step in the clinical pathway model as a set of competing risks in a multi-state survival model to account for this censoring. For example, consider admission to the ward upon presenting to the hospital: individuals can transition from ward-to-discharge, ward-to-death or ward-to-ICU. Individuals in the data who were yet to be observed following any of these were then considered to be censored. We utilised a mixture distribution framework, which estimates a multinomial distribution describing the probability that each given transition will occur, then conditional on each probability of transition, subsequently estimates the corresponding parametric distribution describing the length of stay [13]. This mixture distribution approach produces time-to-event and transition probability results that are more straightforward to interpret and to utilise in further modelling work in comparison to the more typical cause-specific hazards approach to analysing competing-risk survival data [14].

Model estimates were produced using the flexsurv package [14] in R [15], with mixture distribution fits produced across each compartment (ward, ICU, post-ICU-ward). Estimates were stratified by age groups as appropriate to the sample size available for fitting (i.e., broader age grouping used where sample size was too low to produce reliable estimates). The distributions of lengths of stay were modelled as gamma distributions across all transitions, with shape and rate parameters varying by age group in the ward compartment, but with only the shape parameter varying by age group in the ICU and post-ICU-ward compartments (due to limited sample size). To improve model fit for the ward-to-death and post-ICU-to-death pathways, fixed probabilities of transition were specified according to the non-parametric Aalen-Johansen estimates. Cumulative survival probability plots were produced across each pathway, comparing the parametric gamma distribution fits with non-parametric Aalen-Johansen estimates to visually assess goodness-of-fit (Figure 7, 8). All code required to reproduce the results is available at https://github.com/ruarai/los_analysis_competing_risks/. Full tables of estimated parameters are available for download at https://osf.io/uns7v/?view_only=2b4eff42e4b846f0a7768bdb183fc4db.

## Results

We estimated that the mean length of stay for patients hospitalised during the mixed Delta and Omicron epidemic period (15 December 2021 to 7 February 2022) was reduced by a factor of approximately half across most clinical pathways compared to the Delta-only period (1 July 2021 to 14 December 2021) (Figure 4, Table 1). Figure 4 shows that the estimated mean lengths of stay across all age groups and transitions were shorter again when restricting the analysis to cases hospitalised within the Hunter New England local health district (LHD), where it was reported that Omicron was almost certainly the only circulating variant. As has been observed previously, the increase in mean length of stay by age is present for each period for stays in the ward (e.g., [11]).

**Figure 4:**
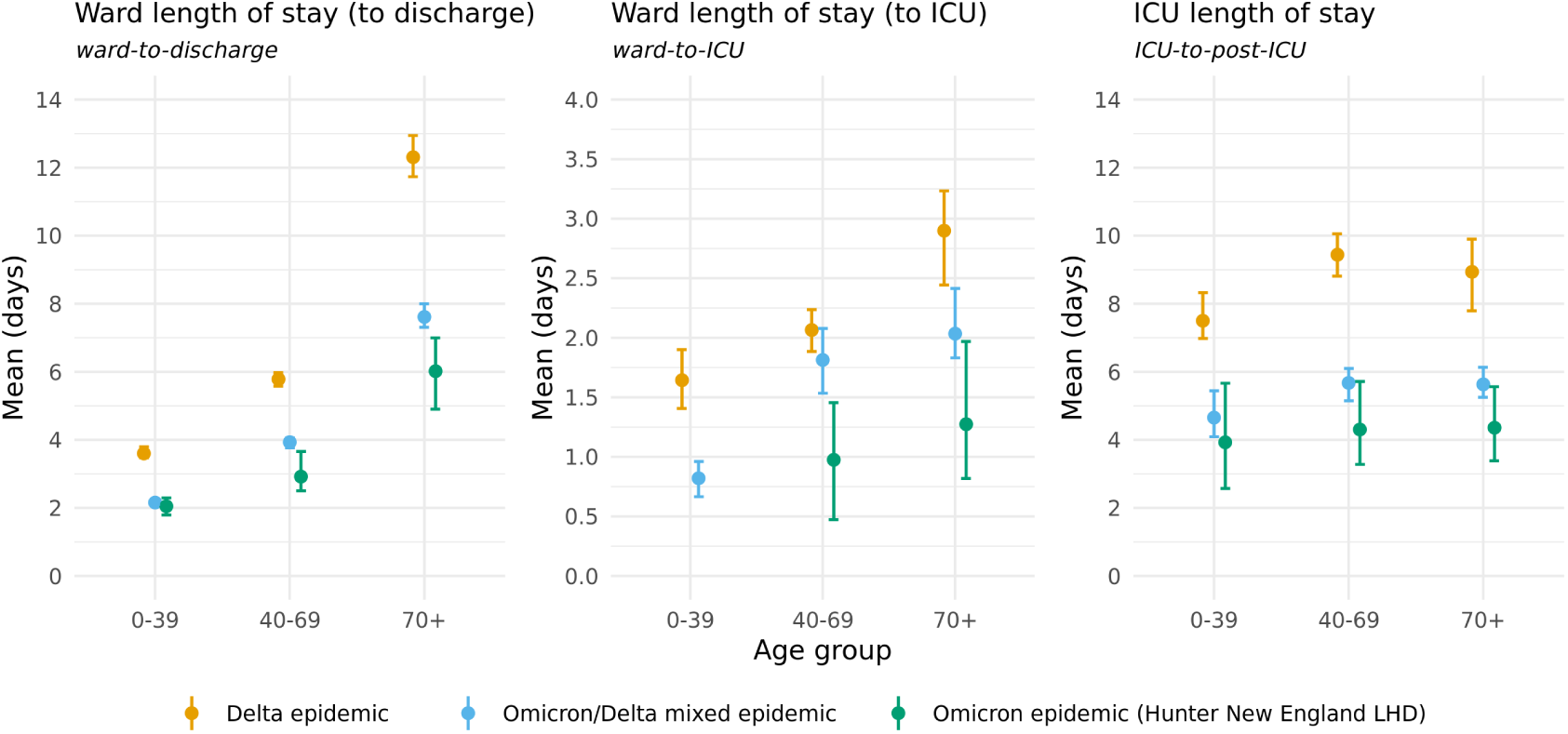
Modelled mean length of stays for the ward-to-discharge, ward-to-ICU and ICU-to-post-ICU pathways. Means and 95% confidence intervals are shown. Note that the y-axis differs between panels. Delta estimates are produced over individuals admitted to hospital between 1 July 2021 and 14 December 2021. Omicron and mixed-Omicron-Delta estimates are produced over individuals admitted to hospital between 15 December 2021 and 7 February 2022. Length of stay estimates for the ICU pathway do not include patients who are recorded as having been discharged directly from the ICU (i.e. not including the direct ICU-to-discharge pathway). Estimates from the Hunter New England Omicron dataset are not displayed where sample size was insufficient to produce a model fit.

**Table 1:**
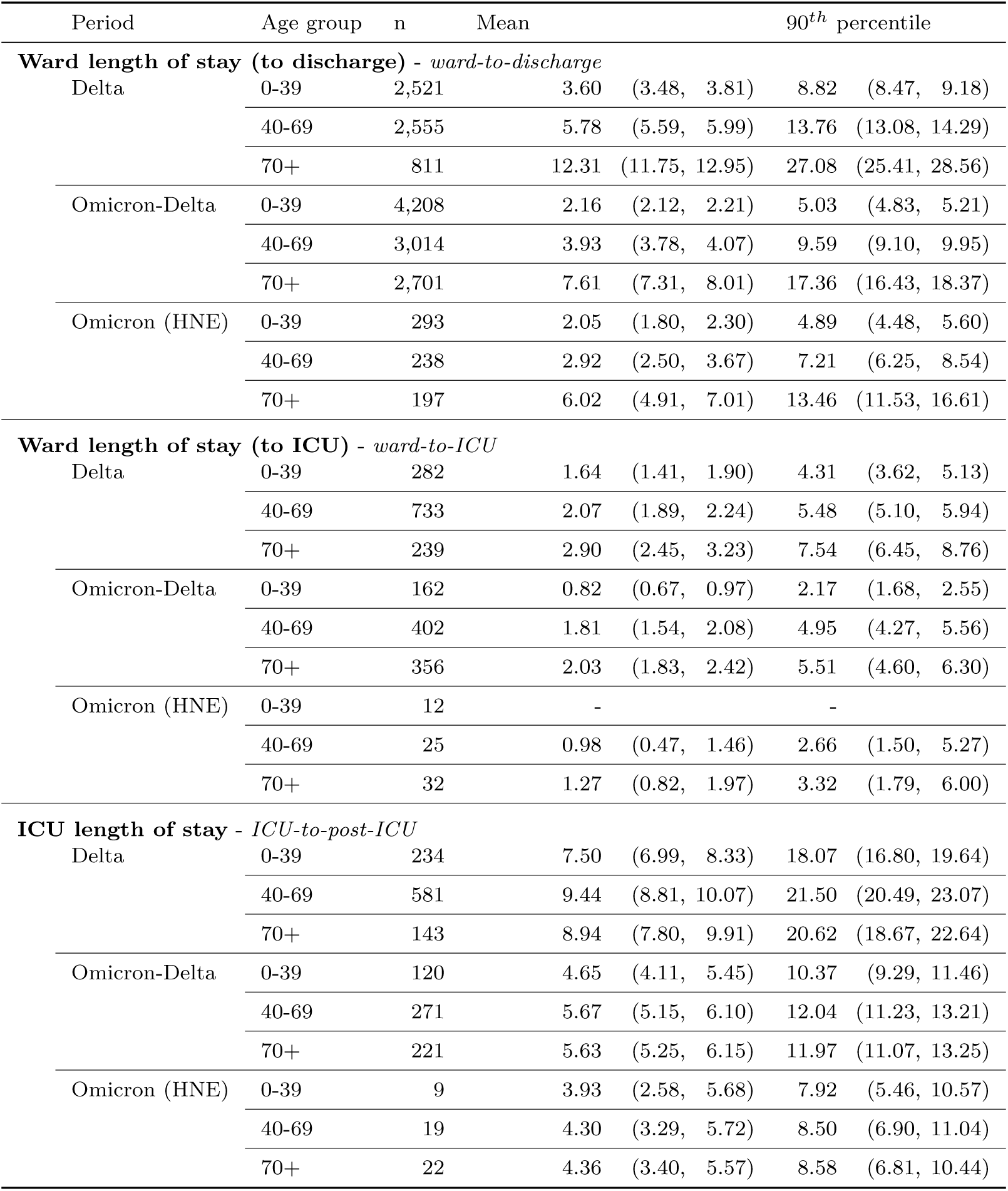
Number of patients observed (n), modelled mean and 90th percentile length of stay (95% confidence intervals) across the ward-to-discharge, ward-to-ICU and ICU-to-post-ICU pathways for all age groups. Delta estimates are produced over individuals admitted to hospital between 1 July 2021 and 14 December 2021, Omicron and mixed-Omicron-Delta estimates are produced over individuals admitted to hospital between 15 December 2021 and 7 February 2022. For each of the Delta, Omicron-Delta, and Omicron (HNE) analyses, there were (3, 1128, 6) censored individuals in the ward, (0, 4, 114) in the ICU, and (1, 135, 10) in the post-ICU ward, respectively.

Table 1 contains the estimated mean length of stay and upper 90^*th*^-percentiles (with 95% confidence intervals) for each transition by age group (as described above) and during each of the Delta, Omicron-Delta and Omicron epidemics. Table 2 in the Supplementary Material reports the corresponding scale and shape parameters, their 95% confidence intervals and the correlation between these parameter estimates, and probabilities of transitions between model compartments, such that they may inform further modelling work. Figure 6 further displays the correlation between the shape and scale parameter estimates via a bootstrap approach.

**Table 2:** Mixed Omicron-Delta epidemic period length of stay and transition probability parameter estimate means and 95% confidence intervals, with sample size (n) and correlation (Cor.) between the natural logarithms of the estimated shape and scale parameters. Note that ward-to-death and post-ICU-to-death probability estimates are produced using the Aalen-Johansen non-parametric estimator and are not displayed. * Note that the probabilities reported here are estimated as described in the Methods section in the main text, and are not adjusted for co-morbidities, vaccination status, prior infection with SARS-CoV-2, sex, or other variables associated with severity of disease. They do not represent a formal analysis of Omicron severity — they represent estimates of the observed transition probabilities during the mixed Omicron-Delta epidemic in the largely previously uninfected study population. We caution against generalising these values to other settings, particularly with different demographics, vaccination coverage, or prevalence of comorbidities, or assuming they would remain constant in other time-periods in the study population.

| Pathway | Age | n | Scale | Shape | Cor. | Probability* |
| --- | --- | --- | --- | --- | --- | --- |
| ward-to-discharge | 0-39 | 4208 | 2.27 [ 2.15, 2.39] | 0.95 [ 0.91, 0.98] | -0.85 | 0.96 [ 0.95, 0.96] |
|  | 40-69 | 3014 | 4.97 [ 4.65, 5.29] | 0.79 [ 0.76, 0.83] | -0.82 | 0.87 [ 0.86, 0.88] |
|  | 70+ | 2701 | 7.32 [ 6.76, 7.95] | 1.04 [ 0.99, 1.09] | -0.87 | 0.78 [ 0.77, 0.80] |
| ward-to-ICU | 0-39 | 162 | 1.48 [ 1.09, 1.99] | 0.56 [ 0.46, 0.67] | -0.68 | 0.04 [ 0.03, 0.04] |
|  | 40-69 | 402 | 3.79 [ 3.07, 4.58] | 0.48 [ 0.43, 0.54] | -0.60 | 0.11 [ 0.10, 0.12] |
|  | 70+ | 356 | 4.09 [ 3.28, 5.01] | 0.50 [ 0.44, 0.57] | -0.64 | 0.09 [ 0.08, 0.10] |
| ward-to-death | all | 289 | 41.51 [ 32.06, 52.43] | 0.82 [ 0.71, 0.92] | -0.82 | – [– , –] |
| ICU-to-discharge | 0-69 | 62 | 10.69 [ 6.10, 17.06] | 0.69 [ 0.52, 0.93] | -0.65 | 0.13 [ 0.10, 0.16] |
|  | 70+ | 25 | 10.69 [ 6.10, 17.06] | 0.69 [ 0.45, 1.01] | -0.47 | 0.08 [ 0.06, 0.12] |
| ICU-to-death | 0-69 | 48 | 20.56 [ 13.69, 29.35] | 0.97 [ 0.74, 1.26] | -0.66 | 0.13 [ 0.10, 0.17] |
|  | 70+ | 55 | 20.56 [ 13.69, 29.35] | 0.87 [ 0.67, 1.11] | -0.67 | 0.23 [ 0.19, 0.28] |
| ICU-to-post-ICU | 0-69 | 391 | 4.14 [ 3.58, 4.74] | 1.30 [ 1.17, 1.45] | -0.79 | 0.74 [ 0.70, 0.78] |
|  | 70+ | 221 | 4.14 [ 3.58, 4.74] | 1.37 [ 1.21, 1.55] | -0.67 | 0.69 [ 0.63, 0.74] |
| post-ICU-to-discharge | 0-39 | 110 | 4.36 [ 3.60, 5.22] | 1.12 [ 0.94, 1.32] | -0.62 | 0.97 [ 0.89, 0.99] |
|  | 40-69 | 209 | 4.36 [ 3.60, 5.22] | 1.62 [ 1.40, 1.87] | -0.77 | 0.93 [ 0.86, 0.97] |
|  | 70+ | 134 | 4.36 [ 3.60, 5.22] | 2.00 [ 1.70, 2.31] | -0.76 | 0.77 [ 0.69, 0.84] |
| post-ICU-to-death | all | 24 | 15.17 [ 1.60, 67.15] | 0.95 [ 0.46, 1.78] | -0.88 | – [– , –] |

The probabilities reported in Table 2 are estimated as described above, and are not adjusted for co-morbidities, vaccination status, prior infection with SARS-CoV-2, sex, or other variables associated with severity of disease (e.g., [16, 17]). These are estimates of the observed transition probabilities during the mixed Omicron-Delta epidemic in the largely previously uninfected study population over a specific time period — they do not represent a formal severity analysis. We caution against generalising these values to other settings, particularly with different demographics, vaccination coverage, models of clinical care, or prevalence of comorbidities, or assuming that these probabilities remain constant in other time-periods in the study population.

Figure 5 shows the estimated length of stay distributions compared to the Aalen-Johansen estimates. These figures indicate that the model provides a reasonable fit to the data. The change in scale (y-axis) indicates the probability of discharge directly from ward for all age groups is higher for the Omicron-Delta and Omicron periods, compared to the Delta period. Conversely, the probability of admission to ICU from ward is substantially lower for all age groups in the Omicron-Delta and Omicron periods, compared to the Delta period. The probabilities of admission to ward following ICU are similar across each age group, though note that these estimates are based on small numbers for the Omicron period.

**Figure 5:**
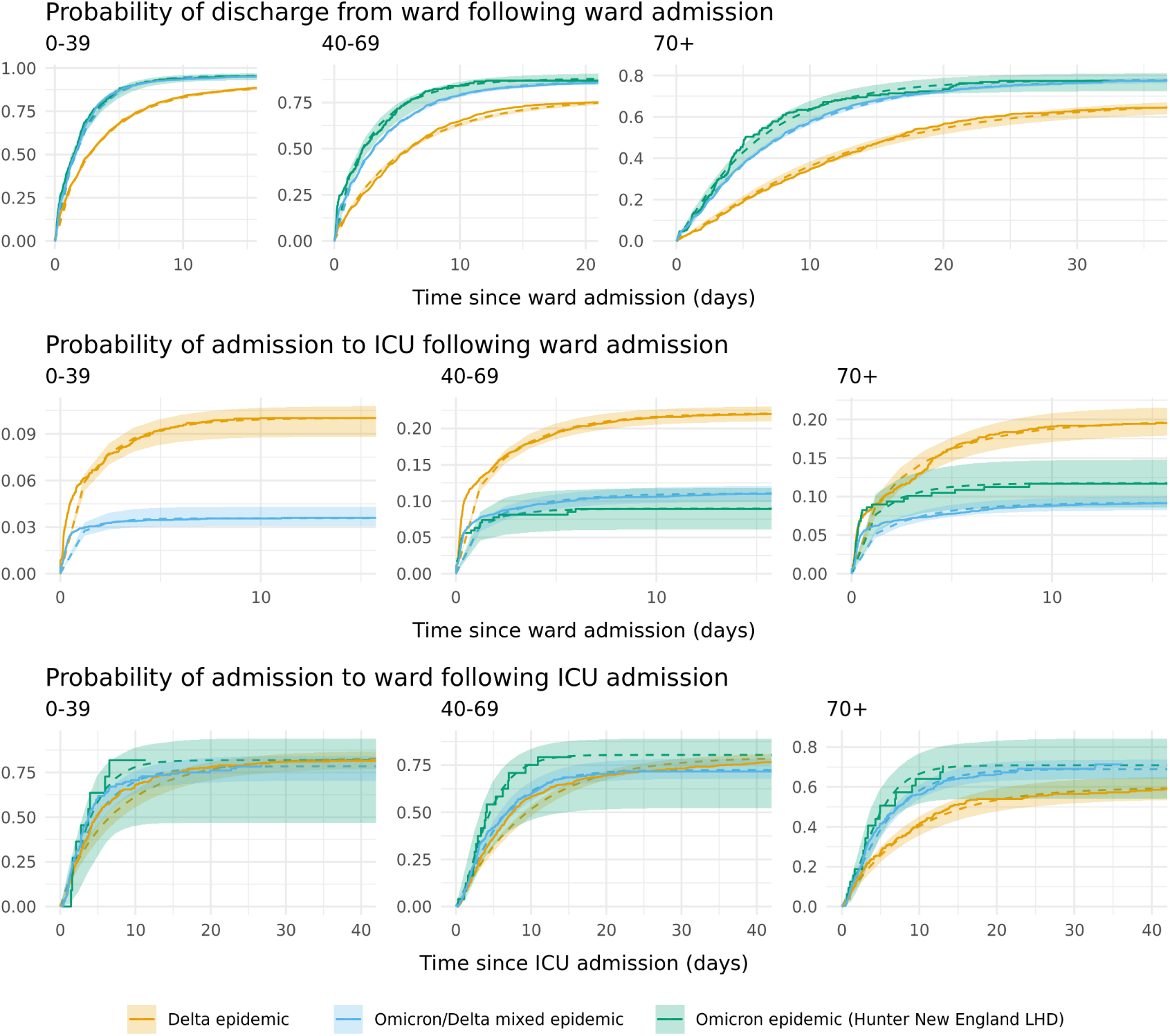
Cumulative survival probabilities of individuals in the ward-to-discharge, ward-to-ICU and ICU-to-post-ICU pathways, across epidemic periods and age groups. Solid lines represent observed data via Aalen-Johansen non-parametric estimates. Dashed lines and shaded regions represent the fit mixture distribution model means and 95% confidence intervals respectively. Note that y- and x-axis extents differ across both pathways and age groups. Delta estimates are produced over individuals admitted to hospital between 1 July 2021 and 14 December 2021, Omicron and mixed-Omicron-Delta estimates are produced over individuals admitted to hospital between 15 December 2021 and 7 February 2022. Estimates for the ICU-to-post-ICU pathway could not be produced from the Hunter New England Omicron epidemic in the ward-to-ICU pathway for age group 0-39 due to limited sample size.

**Figure 6:**
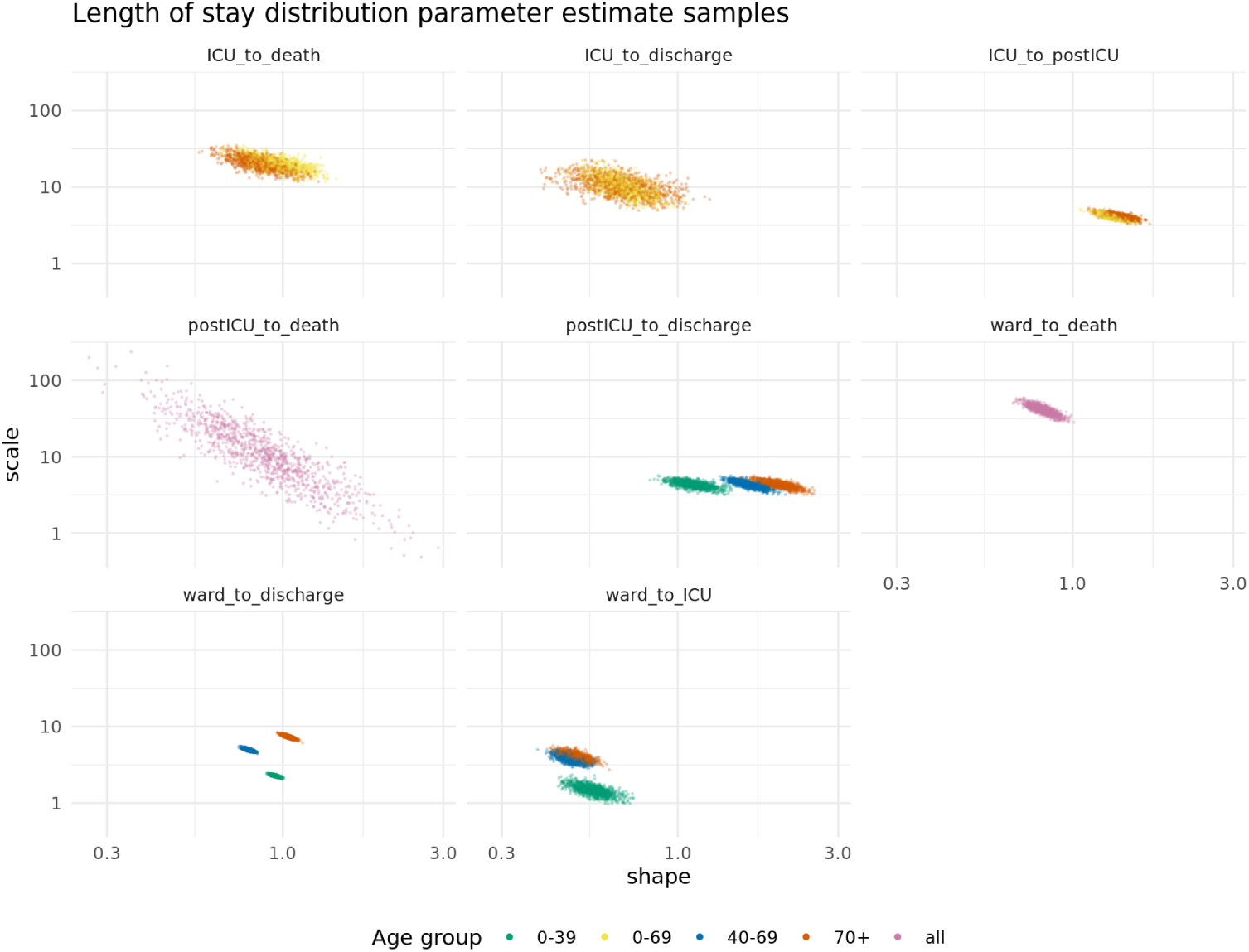
Mixed Omicron-Delta epidemic period length of stay parameter samples.

**Figure 7:**
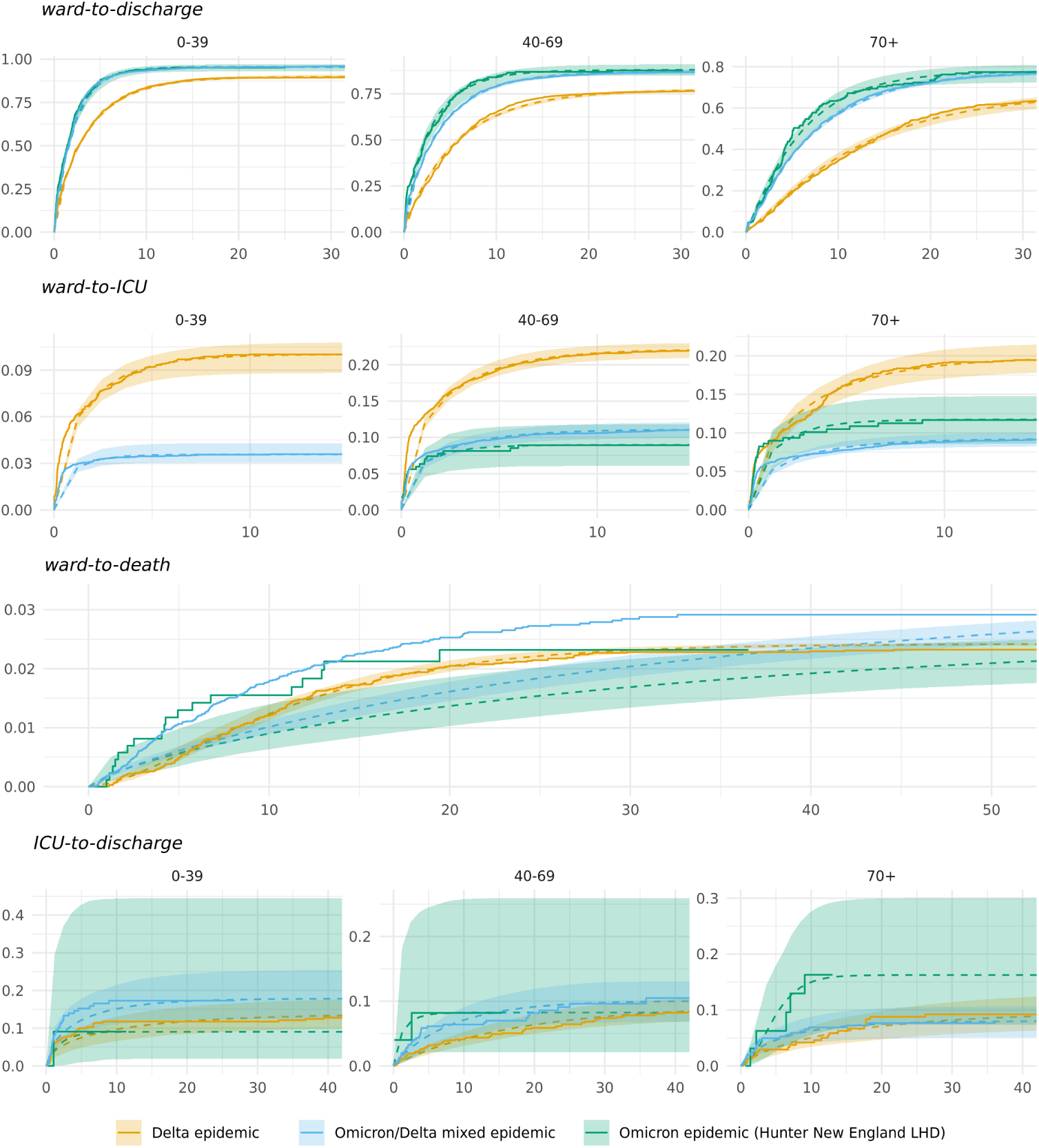
Cumulative survival probabilities of individuals by each pathway, across different epidemic periods and age groups. Solid lines represent observed data via Aalen-Johansen non-parametric estimates. Dashed lines and shaded regions represent the fit mixture distribution model means and 95% confidence intervals respectively. Delta estimates are produced over individuals admitted to hospital between 1 July 2021 and 14 December 2021, Omicron and mixed-Omicron-Delta estimates are produced over individuals admitted to hospital between 15 December 2021 and 7 February 2022. Estimates for the ICU-to-post-ICU pathway could not be produced from the Hunter New England Omicron epidemic in the ICU and post-ICU pathways due to limited sample counts in the data.

**Figure 8:**
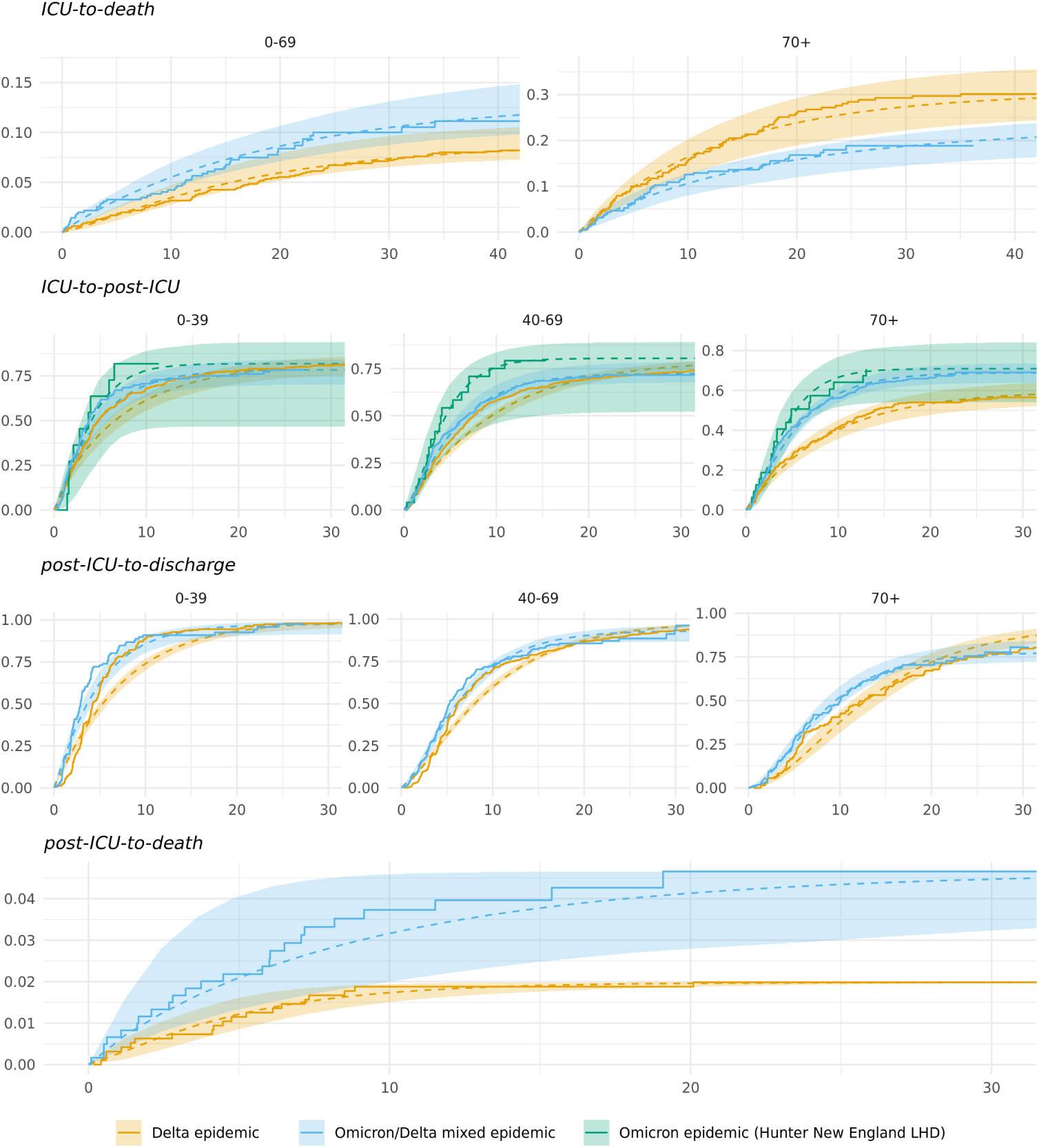
Cumulative survival probabilities of individuals by each pathway, across different epidemic periods and age groups. Solid lines represent observed data via Aalen-Johansen non-parametric estimates. Dashed lines and shaded regions represent the fit mixture distribution model means and 95% confidence intervals respectively. Delta estimates are produced over individuals admitted to hospital between 1 July 2021 and 14 December 2021, Omicron and mixed-Omicron-Delta estimates are produced over individuals admitted to hospital between 15 December 2021 and 7 February 2022. Estimates for the ICU-to-post-ICU pathway could not be produced from the Hunter New England Omicron epidemic in the ICU and post-ICU pathways due to limited sample counts in the data.

The sensitivity analysis shown in Figure 9 indicates that the estimated mean lengths of stay are robust to the data filtering steps implemented. The only differences of note were in the 70+ age group for the ward-to-discharge pathway during the Delta period. Filtering individuals with symptoms after admission results in a substantial reduction in mean length of stay for this age group, from approximately 20 to 12.5 days. This is suggestive of a number of individuals in this age group having long hospital stays with infection incidental to their stay, which our multi-state model does not consider.

**Figure 9:**
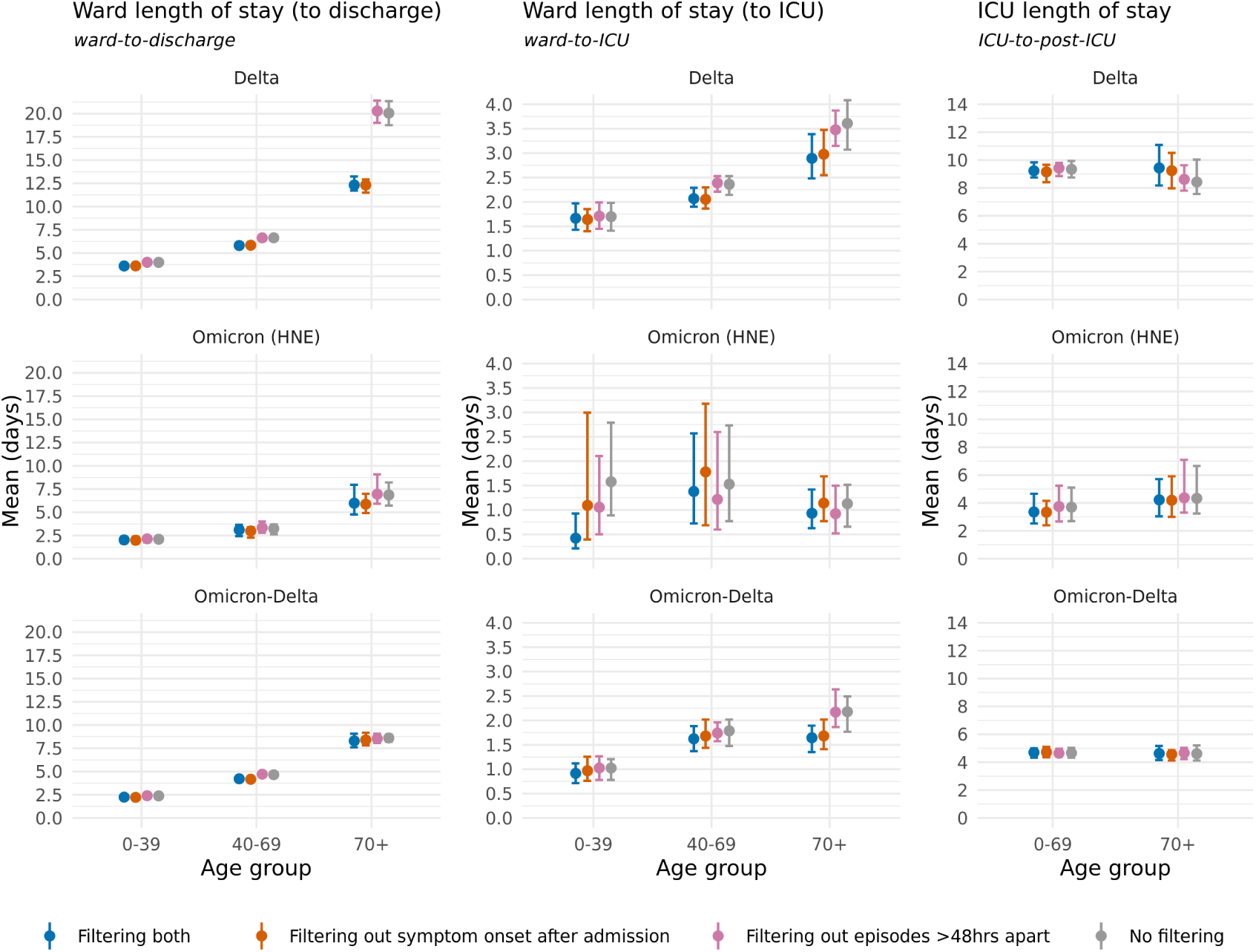
Sensitivity analysis across differing degrees of filtering during construction of the clinical datasets. Estimated length of stay means and 95% confidence intervals shown. For the ‘No filtering’ and ‘Filtering out symptom onset after admission’ scenarios, individuals with episodes greater than 5 days apart were still removed. Data as of 2022-01-25.

## Discussion

The results presented from hospitalised patients in New South Wales, Australia, indicate a reduced mean length of stay of the Omicron variant of SARS-CoV-2 compared to the Delta variant. The observed survival curves and 90% quantile estimates of length of stays are similarly reduced, showing that the overall distributions of length of stay are reduced, and not just the central tendencies. The multistate modelling framework implemented here is a robust approach to estimating hospital lengths of stay during an ongoing epidemic [12]. The data used in these analyses are relatively complete, with at most 11% of data informing each transition and length of stay being censored. Appropriate characterisation of the demand on ward and ICU resources, coupled with estimates of probability of hospitalisation, are paramount to appropriately forecast clinical demand for the Omicron variant and support policy decisions.

The reduced length of stay estimates we have produced are consistent with reports of Omicron-dominated epidemics in other settings internationally, for example: An analysis of a private hospital network in South Africa across 2,351 Omicron patients and 6,342 Delta patients showed a reduction in median total length of stay from approximately 7 − 8 days in the Delta epidemic wave to 3 days in the Omicron epidemic [18]; an analysis of patients within a Houston healthcare network showed a reduction in median total length of stay from 5.4 days for Delta patients to 3.0 days for Omicron patients [19], and; an analysis of a Portuguese cohort reported a 4 day reduction in the mean length of hospital stay between Delta and Omicron [20].

We found that, across all of the Delta, mixed Delta-Omicron and Omicron Hunter New England analyses, the length of stay in ward increased with age group, a pattern which has been reported elsewhere [16, 21]. However, this trend is not observed to the same extent in the ICU pathways, with only moderate increases in the 40-69 and 70+ age-groups compared to those aged 0-39. There is no clear mechanistic explanation for this difference in trend between ward and ICU, though it may be a form of selection bias where those who are admitted to the ICU are triaged conditional on them having a greater chance of benefiting (e.g. more likely to survive, may recover at a greater rate), especially where ICU resources are relatively scarce [22]. However, there have been substantial differences in the rollout of vaccination by age group that may confound this, with a general prioritisation of those at most risk of severe disease for both the first 2 doses and the 3rd dose. Furthermore, another explanation may exist in that given the probability of severe outcomes increases with age, there may be an impetus to keep older individuals in a ward bed for longer periods of observation, which would not be expected to similarly occur in the ICU.

Although there is now strong evidence that Omicron leads to less severe disease at both the individual and population level relative to Delta [23, 18, 19, 20], other factors that influence length of stay must be considered when interpreting the results presented here. At the time of analysis, ward bed occupancy in New South Wales for the mixed-Delta-Omicron epidemic had reached more than double the peak occupancy reported during the Delta-only epidemic (Figure 1). While we do not have any data on the availability of beds at specific hospitals, prior research indicates that increased pressure on health systems leads to self-regulation where earlier discharges will be more likely [24], which would be reflected in our results as a shorter length of stay.

It has been suggested that vaccination may result in a reduced length of stay [25], though evidence in this regard is limited. The reduced length of stay observed between the two epidemic periods could be a result of the changes in vaccination coverage. At the start of the Delta epidemic period (1 July 2021), 6.5% of the total New South Wales population had received two doses of a COVID-19 vaccine, increasing to 52.4% mid-period (1 October 2021), and reaching a coverage of 75.0% by 14 December 2021. The booster/third dose program commenced around late-November 2021, with 24.5% of the total population having received a third/booster dose by 7 February 2022. Population immunity from prior infection at the time of Omicron emergence in New South Wales was negligible, with the count of cumulative confirmed cases reaching only approximately 1% of the population size (with very high levels of case ascertainment achieved through intensive contact tracing).

The reduced length of stay for patients during an epidemic period dominated by Omicron is concordant with the consensus that the variant has a relative reduction in severity compared to the Delta strain [23]. In this work we do not present estimates of other descriptors of severity, such as the probability of hospitalisation or admission to the ICU, as case ascertainment likely changed substantially throughout the Omicron epidemic period. In the absence of data on case ascertainment (as could be produced through infection or serological surveys [26]) any estimate requiring a case or infection count denominator is likely to not be generalisable outside of its immediate context. Despite these challenges, these estimates of the hospital length of stay provide crucial evidence to support ongoing policy planning for current and future outbreaks.

## Data Availability

The analyses performed in the manuscript required access to patient administration data provided through the New South Wales Notifiable Conditions Information Management System (NCIMS), which is not able to be shared under the terms of our data access agreement. For further details on NCIMS, contact the NSW Ministry of Health.

## Ethics Statement

The study was undertaken to inform the New South Wales COVID-19 pandemic response as part of a broader program of public health surveillance carried out under the NSW Public Health Act (2010). The study used routinely collected patient administration data from NSW public hospitals and the NSW Notifiable Conditions Information Management System (NCIMS). Data was supplied by the NSW Ministry of Health, with all records de-identified and securely managed to ensure patient privacy and to ensure the study’s compliance with the National Health and Medical Research Council’s Ethical Considerations in Quality Assurance and Evaluation Activities. The NSW Public Health Act (2010) allows for such release of data to identify and monitor risk factors for diseases and conditions that have a substantial adverse impact on the population and to improve service delivery. Following review, the NSW Ministry of Health determined that this study met that threshold and therefore provided approval for the study to proceed. The project oversight and approval for publication was provided by the NSW Ministry of Health.

## Supplementary Figures

